# Healthcare professionals’ attitudes towards digital health interventions and perspectives on digital health inequalities: a qualitative study

**DOI:** 10.1101/2024.07.06.24310027

**Authors:** Mel Ramasawmy, David Sunkersing, Lydia Poole, Ann Blandford, Paramjit Gill, Kamlesh Khunti, Shivali H Modha, Kiran Patel, Henry WW Potts, Madiha Sajid, Nushrat Khan, Amitava Banerjee

**Affiliations:** Institute of Health Informatics, UCL, 222 Euston Road, London, NW1 2DA, UK; Department of Psychological Interventions, School of Psychology, University of Surrey, Guildford, Surrey, UK; UCL Interaction Centre, London, UK; Warwick Medical School, University of Warwick, Coventry, UK; Diabetes Research Centre, Leicester General Hospital, University of Leicester, Leicester, UK; Patient and Public Involvement Representative, DISC Study, London, UK; Institute of Child Health, UCL, London, UK

**Keywords:** Digital health, cardiometabolic disease, minority ethnic, inequalities

## Abstract

**Background:** Use of Digital Health Interventions (DHIs) for the management of cardiometabolic diseases has increased, but may exacerbate existing health inequalities. Healthcare professionals (HCPs) play a vital role in recommending and supporting patients to use these tools. There is a need to understand the role of HCPs in managing the implementation of digital health in communities at risk of health inequalities.

**Objective:** To explore the views of health care professionals about digital health and its impact on health inequalities, focusing on cardiometabolic diseases and the South Asian population in the UK.

**Methods:** Online interviews and focus-groups with HCPs (n=18), exploring participants’ experiences and attitudes towards digital health, perceptions of patients’ barriers and facilitators to use, whether they perceived any populations to be particularly at risk of digital inequalities, and the impact of the COVID-19 pandemic on their practice in relation to digital health. After informed consent, transcription and coding, a reflexive thematic approach was taken to analysis.

**Results:** HCPs recognised the potential benefits of DHIs to improve access and delivery of care and patient outcomes, but described several barriers to successful implementation. HCPs demonstrated a good understanding of the challenges their patients face in relation to wider inequalities, barriers to health behaviours and healthcare access, and digital health. Of particular concern was the impact of increasing reliance of digital interventions in health care on the exclusion of some patient groups. Participants recommended improvement of the design and implementation of DHIs offered to patients through working with at-risk populations throughout the process. Finally, participants emphasised the importance of ensuring non-digital services remained available to ensure equitable access to health and social care.

**Conclusions:** HCPs described the complexities of delivering care to underserved communities. DHIs were identified as a potential way to improve health outcomes for some, while over-reliance risked exacerbating inequalities. Participant recommendations related to design, implementation, and engaging target populations providing practical examples to address digital health inequalities, such as working with other sectors to take a community approach.

## Background

Digital health interventions (DHIs) have the potential to improve patient knowledge and outcomes, and save health care costs [1–4]. The UK government is investing significantly in digital transformation and innovation [5], and patients can now use their online National Health Service (NHS) account to access NHS and commissioned services such as managing bookings and referrals, accessing healthcare records, and using online pharmacies [6].

Health care professionals (HCPs) play a pivotal role in the introduction and uptake of DHIs. Previous studies have shown variation in the digital health competence of HCPs [7], and highlighted barriers and facilitators to HCP use of digital health, including infrastructure, technical barriers, training, evidence about technology effectiveness, concerns about workload, and individual personal and psychological barriers, such as resistance to change or concerns about losing human interaction [8]. There has also been some exploration of HCPs attitudes and behaviour in relation to apps for specific conditions such as depression [9]; or patient use of wearables [10].

The implementation of DHIs may exacerbate existing health inequalities, for example by age, ethnicity, socio-economic status, and health conditions [11]. There is therefore a need to understand the role of HCPs in managing the implementation of digital health in communities at risk of health inequalities. We aimed to understand HCPs’ perceptions of digital health and health inequalities. We focused on cardiometabolic diseases and the South Asian population in the UK as a case study, as a group that experience poorer cardiometabolic health outcomes and are more likely to experience barriers to digital inclusion [12,13].

## Methods

### Study Design

A qualitative approach encompassing interviews and focus groups was taken. The study received ethical approval from NHS London – Brent Research Ethics Committee (IRAS 261047). Study reporting was completed in line with COREQ guidelines (see Supplementary 1).

### Focus Groups and interviews

Recruitment of HCPs within the UK took place via primary care, professional networks and snowball sampling methods [14], to reach a range of professional groups involved in the care of those with cardiometabolic diseases, or in diverse populations. If participants were not available to attend an online focus group (lasting 1 hour), interviews were offered instead to enable them to participate (lasting 30 minutes to one hour). Interviews took place between April and December 2022.

Before each focus group or interview, participants provided written informed consent, with any queries addressed by the research team. The discussions explored professionals’ experiences and attitudes towards digital health, perceptions of patients’ barriers and facilitators to use, whether they perceived any populations to be particularly at risk of digital inequalities, and the impact of the COVID-19 pandemic on their practice in relation to digital health (full details are provided in Supplementary material 1). Participants were offered a £50 retail voucher for their time.

Interviews were conducted by MR and NK who are both experienced qualitative researchers. Interviews and focus groups were recorded and transcribed verbatim using Microsoft Teams, and were checked and anonymised by the research team afterwards. A reflexive thematic approach was taken to analysis [15]. After familiarisation, transcripts were coded by MR and NK. An iterative process of coding, review and revision of codes was completed, and codes were organised into themes through discussion between MR, NK, and DS.

## Findings

### Participant characteristics

A total of 18 HCPs working across primary, community and secondary care in the UK were recruited (see Table 1). For participants that provided demographic data (n=16, 89%), the mean age was 38 years, and 10 were female (63% of those that provided details). The majority of participants were from a South Asian ethnic background (n=13, 72%), which might reflect interest in the subject and confidence on speaking about ethnic inequalities in health. Digital literacy of participants was not documented.

**Table 1.** Participant demographics.

|  | N (%) |
| --- | --- |
| **Age** |  |
| 20 – 29 | 3 (17%) |
| 30 – 39 | 8 (44%) |
| 40 – 49 | 3 (17%) |
| 50 – 59 | 1 (6%) |
| 60 – 69 | 1 (6%) |
| Not provided | 2 (11%) |
| **Gender** |  |
| Female | 10 (56%) |
| Male | 5 (28%) |
| Other/not provided | 3 (17%) |
| **Ethnicity** |  |
| Asian/Asian British | 13 (72%) |
| White | 3 (17%) |
| Other/not provided | 2 (11%) |
| **Religious beliefs** |  |
| Christian | 1 (6%) |
| Hindu | 6 (33%) |
| Muslim | 4 (22%) |
| Sikh | 2 (11%) |
| None | 3 (17%) |
| Other/not provided | 2 (11%) |
| **Role** |  |
| General Practice | 3 (17%) |
| Pharmacy | 4 (22%) |
| Specialist doctor or nurse - Diabetes | 2 (11%) |
| Specialist doctor or nurse - Cardiology | 1 (6%) |
| Doctor or nurse – Other | 3 (17%) |
| Dietician or Nutritionist | 2 (11%) |
| Other health role | 3 (17%) |

### Healthcare professionals’ attitudes and experiences of DHIs

Participants described various digital health approaches in use in primary and secondary care around monitoring, information provision and appointment or medication administration, with a range of complexities. Specific examples related to CMD monitoring ranged from providing patients with low-cost blood pressure devices, to the NHS ‘Heart failure @ home’ programme [16]. There was a perception of different levels of acceptability of the implementation of digital in health within and between primary and secondary care, different specialities, and within pharmacy where patients often sought additional advice.

Health care professionals highlighted potential benefits of digital approaches in the NHS, such as collecting data that supported consultations, speeding up diagnosis and treatment, and managing waiting lists. They praised the positive impacts of technologies on patient self-management and outcomes, such as continuous glucose monitoring, and the potential for improved communication and follow-up of relevant information, through SMS messages, and links to leaflets, videos and websites.

> *“But understandably, when patients are having appointments with clinicians whereby we only have 10 to 15 minutes to discuss an issue, then we tend to use these particular types of leaflets and videos [sent by text message] as a bit of a supplementation to what we’ve discussed in the consultation*.*” [P5, Pharmacist]*

However, they also highlighted that those benefits were not yet always realised, for example, on a platform designed for patients to undertake and record their own blood pressure readings, one clinician noted that “about 60-70% of the time you’ve got to chase them anyway… [it] wasn’t as beneficial as we’d have liked.” [P10, Consultant in Diabetes]. Another reflected on the potential risk of harm, such as patients being given access to their electronic health records without adequate support:

> *“… not only do people look at the record and, you know, start to query what the doctor’s written, or they don’t really understand what the doctor’s written… they’ll Google it and then they’ll see that actually my result is abnormal, [but they are looking at the wrong measurements]” [P18, Junior doctor]*.

Participants described mixed attitudes of patients towards DHIs, some of which were dependent on the type and immediacy of the health condition. For example, patients were described to be happy to have routine appointments over the phone, but would prefer to see a HCP face-to-face for new conditions or where required, such as diabetic foot checks. From our research with patients, we know that this was not happening, even outside of lockdown restrictions [17]. Participants reported that patients with diabetes were very engaged by continuous glucose monitoring, seeing it as preferable to finger pricking. HCPs reported that other interventions such as DAFNE (‘Dose Adjustment For Normal Eating’, an NHS Type 1 diabetes education programme) [18] and digital weight loss programmes had a more mixed uptake and saw high drop-off after referral. This is supported by evaluations of these programmes [19,20].

They noted that patients were often already using smartphones for things of interest to them, such as speaking to family members, and that the increased use of digital applications in non-health contexts had also increased acceptability in health contexts. Several HCPs spoke about how, in contrast to their expectations, patients felt enabled by technology, and wanted to own and use their health data and access the latest technology to help manage their condition.

> *“[With continuous glucose monitoring] - the view was patients weren’t going to be very interested in this - but you know that I think all of our experience it’s the patients are the ones driving it and NHS - we have to catch up!” [P2, Academic Clinician (Nursing)]*

However, they also noted patient concerns about data privacy, and that this overlapped with other hesitancy to engage with healthcare, such as vaccine hesitancy.

> *“… for example, the COVID vaccination program, especially amongst the BAME [Black Asian and Minority Ethnic] population… there was a lot of vaccine hesitancy and that’s quite well documented. And I guess you can quite easily correlate that with some of the interventions that require people to put in personal information. There might be this element of, ‘well, what on earth are they gonna do with this information?’” [P5, Pharmacist]*

There was also a view that patients perceived remote care as of lower quality, or represented HCPs avoiding seeing patients.

> *“They feel like you’re fobbing them off if you give them something online, it might be better, but they feel like clinicians are trying to steer them away from GP practice because they want to save time. They want to save money*.*” [P5, Pharmacist]*

Clinicians noted that an important factor in overcoming this was the patient relationship, reinforcing the use of digital tools and supporting effective use through follow up.

> ***“****I would also want to make sure that it could be followed up effectively… It’s not just something that you give to them and say go away and do this for six weeks. It’s something that you can check in and see that they’re actually following it as… it was intended*.*” [P18, Junior doctor]*

When recommending DHIs to patients, professionals drew on their experience and knowledge, mentioning that there was variability between clinicians. Examples of the types of tools clinicians felt comfortable with recommending included simple commercial apps to improve diet and physical activity levels. Participants spoke about the importance of shared decision-making in promoting adherence, and gave examples of how a decision to use DHIs was often driven by patients’ interest in DHIs and their exposure through friends, family and media.

> *“It’s quite difficult to keep up with [the pace of change in digital health] - you know, often people come to us and say, ‘Well, can I have this device?’ that I’ve never heard of… We’re certainly seeing a bit of push from patients now to [request the latest devices]”* [P4, Consultant in diabetes]

> *“I would volunteer [DHIs] for patients that were struggling or the patients that are saying… ‘Can you advise me on something?’ - and I can say well, I’m familiar with these [commercial diet and exercise apps]. And it is anecdotal feedback from patients, a lot of patients tell me… ‘This app’s good’. So it’s that type of approach - I don’t say actually ‘My advice is you go and use [commercial app] to count your calories*.*’” [P6, GP]*

All participants spoke about how COVID-19 restrictions and the need to deliver care remotely had an impact on digital health offerings, uptake by their patient populations, and openness of the healthcare system to digital tools. Without sufficient NHS services in place for some remote monitoring tools, clinicians made recommendations to use commercial apps to support diagnosis (e.g. for atrial fibrillation). One GP explained how uptake of a commercial platform “dramatically increased” during the pandemic, and how this rapidly changed how they communicated with patients:

> *“[Pre-pandemic] we never sent texts, we never asked for text back from patients, photos and information, we never did video consultations… What we would probably not have done in 5-6 years, we did in a couple of weeks*.*”* [P6, GP]

> *“… it’s not very easy to diagnose atrial fibrillation over the phone. So we just have to rely on patient symptoms. But and also there’s such a now a long wait to have*… *Holter monitoring… So I’ve been recommending to patients because they are obviously feeling quite unwell to buy [commercial DHI]. And that really has made a difference [to diagnosis and initiating treatment]. [P2, Academic Clinician (Nursing)]*

However, participants felt that a lack of evidence about the efficacy of DHIs impacted the advice they were able to give to patients, for example, not being able to make a recommendation directly. They wanted more joined-up commissioning, such as NHS-driven DHI infrastructure, which would enable them to select appropriate tools from a trusted source, know that this was funded in their region, and which would ensure data was shareable between parts of the NHS. This would also benefit patients who were reported to be frustrated by regional variation in the availability of tools, the need to regularly learn new systems, and lack of joined-up data (for example one part of the system not being able to access results of an investigation in another part of the system). Overall, HCPs currently felt restricted, and needed more support, training and information to make recommendations to patients safely.

> *“Everything doesn’t integrate into one system, so you have to kind of go into different logins for different things and that’s really frustrating… trying to get everything into one system. So you can just click one entry point would be great*.. *to simplify it for the health professional, but also for the patient as well*.*”* [P10, Consultant in Diabetes]

> *“For me at the moment, it’s only things that are recommended by NICE. So, yeah. I mean, we’re very hamstrung with the number of patients that we can refer for various technologies and so on.”* [P4, Consultant in diabetes]

### Perceptions of inequalities and intersection with digital exclusion

Health care professionals had a nuanced understanding of the challenges and needs of their patient populations, and the intersecting factors that contributed to health inequalities. This included reflecting on the impact of the cost-of-living crisis, the difficulties of providing care around prevention and management of cardiometabolic diseases in deprived communities, and the lack of resources, such as interpretation, to support patients facing inequalities in access.

> *“[Rare diabetic emergencies] seem to be becoming more prevalent and it’s as a result of COVID and sort of the pressure on people. I think also going back to heating versus eating that is a huge problem, you know, so patients can’t afford bus fares or train fares to come to hospital, and that’s gonna be a big issue, which no one’s really sort of considered. So we roll out 5G. Yeah, great. But no one can - no ones going to be using the technology because they’re afraid of more costs*.*” [P2, Academic Clinician (Nursing)]*

HCPs also discussed the impact of the changing food environment on population health as something beyond their scope. This requires intervention at a policy and local government level [21].

> *“Where the [primary care practice is based]… 20 years ago had a mixture of shops, and now it’s just 90% takeouts… So you know, so one of the biggest battles would be fast food. More than anything else and you don’t know what one can do about that*.*” [P15, GP]*

Participants described inequalities they observed in their areas of work as being more related to social deprivation than specific ethnic or cultural groups. An example of this was that new migrants to the UK (such as those from Eastern Europe who arrived after Brexit) were showing the same patterns of health problems and lack of engagement with healthcare services as previous generations of South Asian migrants.

Health care professionals also reflected on their positionality (their social identities) and how this affected their ability to engage with diverse patients. For example, one South Asian GP, speaking about family dynamics and their role in promoting health behaviour, explained how he can engage the whole family in health changes, using his familiarity with South Asian cultural norms: *“… the children generally are quite involved… and they usually live together so… I also tell them that if their mum or dad is diabetic, then they’re also more likely to have diabetes if it’s the son or the daughter. So then that way it kind of helps to try and improve everyone’s diet altogether so that they’re quite keen on that*” [P6]. He reflected on a recent appointment with another patient, who came in on his own, and “*said he didn’t really want to bother his children to make, like, special food just for him*”, and how in other cultures and family dynamics, he was not able to use that same strategy.

Several participants commented on the intersection between deprivation and health literacy, and how these might interact with potential benefits of digital implementation. For example, one Primary Care Pharmacist explained:

> *“The people that… don’t have so much money. They definitely struggle with, first of all understanding like the diabetes. And then I have to spend a lot more time with them to explain why we need to get [the condition] under control. And I think that’s why they’re less likely to be proactive and you know, want to have these apps and do these things*.*”* [P7, Primary Care Pharmacist]

Some participants described typical factors related to digital exclusion such as age, generation, language spoken, literacy and education, cost and access to devices, as well as other specific groups at risk such as those leaving prison, those with physical barriers (such as arthritis and sight or hearing impairment), and those with learning difficulties. However, others highlighted that widely held perceptions of inequalities did not necessarily match what was observed in practice:

> *“I’ve been quite surprised at, you know, older Bangladeshi diabetic women who come and see me online via ‘Attend Anywhere’, often with maybe one of their relatives helping them out and so on. Where I’ve always felt, actually these are the sort of people that might not want to engage online, but actually I’ve been very pleasantly surprised…”* [P4, Consultant in diabetes]

### The impact of the digital divide in health care practice

HCP experiences during COVID-19 provided a useful example to reflect on the potential impact of digital services on health inequalities:

> *“*… *at one point our weight loss and diabetes prevention services were purely digital and I only had that to offer and that really made me worry that I’m only giving these people one option and it might not be the appropriate one*.*” [P8, GP]*

Participants shared their observations of significant factors relating to digital health inequalities in practice; less focused on age and ethnicity, and more related to individuals’ digital skills, language skills, and trust and familiarity with DHIs and the healthcare system. Additionally, cost and lack of privacy through use of a shared device impacted DHI uptake. HCPs spoke about how existing pressures, such as short appointment times, made it difficult to assess or provide patients with appropriate information about digital.

> *“Because I think we’re kind of expected to assess people’s digital literacy or their preference before we refer them or suggest. But often you don’t either have time, or you can’t - you just kind of have to get on and make the suggestion or make the referral*.*” [P8, GP]*

Participants felt that both digital and non-digital interventions had the potential to exacerbate existing inequalities. For example, lack of tailoring of advice and guidance to different cultural groups, and lack of information about appropriate community resources such as social prescribing, excluded some groups from benefiting. From the opposite perspective, it was felt that those who benefitted the most from digital were those who already took positive action in relation to their health:

> *“The people I would see in clinic would be the ones who would probably like to engage a bit more and so you would get them [using DHIs]. The ones who wouldn’t turn up… perhaps you would miss a lot of them and that would be a lot more minorities*.*”* [P10, Consultant in diabetes]

While it was felt that service commissioners were recognising the higher risk of health inequalities experienced by some ethnic minority patients, the lack of data around DHI implementation, uptake and use made it difficult to understand the full picture of inequalities.

> *“I do wonder if there’s something around intersectionality… there are other challenges that people face within health, within healthcare… it’s very rare that somebody… has only one challenge… So if you overlay disability, and you know, and learning difficulties and other things into the mix, and I don’t know that there’s been enough data captured - there may be subsets of people that are disadvantaged or missed, it may just have been that they’ve not been offered. So we don’t have any data because inadvertently they’ve not used it, and I don’t know if that data is captured so that anybody knows – because you don’t know what you don’t know, do you?”* [P3, Pharmacist]

Overall, participants showed both concern and optimism about the potential of DHIs in relation to inequalities. A key concern was that over time, services would rely more routinely on digital, reducing the quality of care, and excluding some demographic groups. Others suggested that digital approaches could improve access to health care for some people who would struggle to attend, such as those who found it difficult to leave the house, as well as helping overcome language barriers through videos rather than written resources. Participants also shared some examples of good practice to engage their local population in DHIs and programmes around diabetes management.

> *“We’ve been running [structured diabetes education] online via teams, and*… *I had long conversations with our education team saying you know, are we really going to run this this way and is this the only way we’re going to offer education now? And you know, what about the people who are going to miss out? And again the engagement with education has been really, really positive amongst our South Asians… amongst groups that I wouldn’t have assumed would be very [IT familiar]… What I’m slightly worried about is that it will end up being the only offer that we have*.*”* [P4, Consultant in diabetes]

> *“… Desmond, which is a diabetes education support program. We’ve had a local voluntary service provide that in Urdu and Punjabi as well. Previously, we couldn’t refer to the structured diabetes education program because it was only delivered in English. So for a lot of our patients, that was no good. That’s changed*.*” [P6, GP]*

### Healthcare Professional recommendations for equitable uptake of DHIs

Participants made recommendations related to the equitable uptake of DHIs in three areas: design, implementation, and engaging populations experiencing or at risk of health inequalities through the process. Recommendations around design focussed on improving accessibility of DHIs for patients with a range of access needs, such as using diagrams, simple language, and audio and video options, and improving cultural appropriateness of content (e.g., around food information in healthy living interventions). They highlighted that some systems charge to add additional languages, and while this might be too expensive at an NHS Trust level, could be affordable at a national level. They also drew attention to the need to improve communication of legal, data protection and permissions information in a way that was understandable to the public. To maintain patient interest, the DHI offer should be tailored to individual interest, and the intervention should not be too complex or time-consuming.

> *“It helps to be culturally specific*… *And by that what we mean is - talking in that cultural language. So perhaps using certain words or using certain examples of foods, not just translating things, but it’s much deeper than that*.*” [P16, Dietician]*

Participants recommended the use of low-tech solutions, such as SMS messaging, as a ‘universal approach’, although it was emphasised that DHIs should only be part of a range of options and non-digital services were essential to reach everyone. To support implementation of DHIs more generally, clinicians wanted to see more evidence-based recommendations from trusted organisations such as national charities and commissioning from the NHS, and education and support for health care professionals, including those in pharmacy and new primary and social care roles, to enable them to support patients to select and use DHIs for their health.

Participants felt that there were opportunities to bridge the digital divide. They recommended engaging with patients and the community to understand digital access and literacy needs, and working with existing community structures. This included: drawing on family support to introduce virtual consultations or DHIs, particularly where younger generations had healthcare training; using community interest in health as an information network; providing information via cultural media; and working with local champions. They also highlighted the need to be sensitive to the expectations of the patient population in relation to the role of different parts of the clinical team, and providing education to improve uptake of appointments with the complement of health care professions. To support continued use and benefit from DHIs, participants highlighted the need to think holistically and engage the community to address the facilitators and barriers of the behaviour change approach, rather than just the DHI. They emphasised the need for follow-up and for joint working between primary care, pharmacy, local government, and voluntary organisations to deliver equitable care.

> *“I think health care professionals or community leaders recommending them and supporting them to use it can help with ongoing use as well. So having that kind of check in with people to ask, are you still using it? How are you finding it?” [P8, GP]*

> *“A big part [of supporting people to engage or download their first app] would be played by sort of voluntary and tertiary sector organizations who see these patients within the community, who are trusted by the community, and who are saying ‘Actually, let’s do an educational session…’ … I think some of it would be us in the community where we’ve got… social prescribing link workers, care coordinators - so an element of their role would be to increase engagement with IT solutions, to support self-care and management of chronic conditions*.*” [P6, GP]*

## Discussion

### Principal findings

HCPs appreciated the potential benefits of DHIs to improve access and delivery of care and patient outcomes. Barriers to implementing DHIs in practice included a need for a repository of trusted DHIs, lack of time to introduce and support DHIs to patients, and need for additional training and support. Secondly, HCPs had a good understanding of the challenges their patients faced in relation to wider inequalities, barriers to health behaviours and healthcare access, and digital health. They were concerned that over-reliance on digital interventions within the healthcare system may exacerbate existing inequalities. HCPs identified that groups that are particularly at risk of digital exclusion include those experiencing deprivation, individuals who did not speak English and/or with low literacy, people with learning difficulties, and those with physical impairments that might impact use of particular tools, e.g., sight, hearing, and arthritis. Third, participants made recommendations about how the health system can improve the digital offer, through design, implementation approach and engaging populations experiencing or at risk of health inequalities. Finally, participants emphasised the importance of ensuring non-digital services remained available to ensure equitable access to care.

### Comparison with prior work

Previous studies looking at HCP views on acceptability of the implementation of digital in health have also reported ambivalence, and described both opportunities, for example, increased patient self-management, and concerns, such as around usability, privacy and cost [10]. Common barriers to adoption include individual factors (such as confidence prescribing digital interventions), impact on practice (e.g. time and resource implications), and intervention factors (including lack of evidence for effectiveness, and security concerns) [9,22].

Clinicians also reported infrastructure barriers, suggesting that centralised commissioning would provide assurance, and address additional costs associated with improving accessibility of DHIs. Centralised systems for DHI reimbursement are in place to some extent across Europe[23], but evaluation of the DiGA “app on prescription system” in Germany suggests that there is an emerging divide in DHI uptake [24].

Clinicians reflected both on the enabling impact of the pandemic on both patient and healthcare system openness towards digital health, and its potential impact in worsening inequalities in care access and outcomes. For example, some patients were able to use commercially available apps to support diagnosis of atrial fibrillation. Other research at this time described COVID-19 as a destabilising experience for healthcare providers, and noted that there had been a lack of cultural change to deal with the introduction of telehealth [25].

HCPs in our study discussed how their previous perceptions of who might use DHIs was challenged by the uptake during this time, particularly in relation to older adults. Previous work has highlighted that when HCPs hold stigmatising attitudes about ageing, this can influence use and adoption of DHIs [26], and that the gap between willingness to use and recommendations from HCPs increase with age [27]; this suggests that providing education and support to HCPs to recommend DHIs to a wider range of people may increase uptake in those that might benefit.

In addition to reducing inequalities in how DHIs are offered, participants also suggested that digital approaches might enable more accessible care, for example, through the use of video rather than written sources, reducing the need for travel for those who had financial and health barriers to in-person access, and being more practical for those who might not be able to get time off work. Other studies have also highlighted how DHIs might improve provision of culturally sensitive information, for example HCPs providing food advice to women from diverse backgrounds with gestational diabetes found a culturally sensitive DHI could fill gaps in their knowledge about other food cultures [28]; and others have suggested that advances in artificial intelligence could improve health information access for linguistically diverse populations, including through real-time translation [29].

### Limitations

The majority of participants in the study were from a South Asian ethnic background (n=13, 72%). This may partly reflect the composition of the clinical workforce (42% from black and minority ethnic backgrounds in 2020) [30], and interest or confidence on speaking about ethnic inequalities in digital health. One focus group participant was concerned about using incorrect phrasing and unintentionally causing offence. A Royal College of Physicians’ report on addressing health inequalities in practice found that 67% of clinicians feel they had not received enough training, and only 31% felt confident in their ability to talk to patients about the impact of inequalities on their health [31]. Similarly, a Health Education England review found that many NHS staff reported no training for digital transformation [32]. We addressed this through creating a supportive space for discussion, and offering participants individual interviews if they preferred.

Participant experience with digital cardiometabolic interventions also varied by role; for example, those working in secondary care had more experience with tools such as remote monitoring of atrial fibrillation or blood glucose, while those in primary care spoke about tools to do with lifestyle change, information provision and access to services.

While this study focussed on South Asians, as the largest minority ethnic group in the UK [33], the findings highlighted that approaching the question of DHI solutions via ‘ethnicity’ or other broad social groups was not always considered suitable for real-life practice. Participants spoke instead about barriers to access experienced by individuals (such as literacy, financial, learning disabilities, and physical impairments that could impair smartphone use), or about social barriers to engagement with healthcare, such as language and culture in specific communities in their area. This is reflected in the recommendations on improving DHI design and implementation to improve accessibility and utility for all.

## Conclusion

This study describes HCP perspectives on digital health inequalities with a focus on cardiometabolic diseases. HCPs described the complexities of delivering care to underserved communities, and the potential for digital approaches to both address and exacerbate inequalities. Participants provided recommendations related to design, implementation, and engaging target populations, providing practical examples to address digital health inequalities. Particular emphasis was given to the need for better NHS evaluation and commissioning to support HCPs to utilise DHIs in practice.

## Data Availability

The data that support the findings of this study are available from the corresponding author, MR, upon reasonable request.

## Acknowledgements

The authors were funded by the National Institute of Health Research (NIHR; NIHR200937). The funding source played no role in the design of the study; collection, analysis, and interpretation of data; writing of the report; and decision to submit the paper for publication.

PG was supported by the NIHR Applied Research Collaborations West Midland and is an NIHR Senior Investigator. The views expressed in this publication are those of the authors and not necessarily those of the NIHR or the UK Department of Health and Social Care.

KK is supported by the NIHR Applied Research Collaboration East Midlands and NIHR Leicester Biomedical Research Centre.

LP is supported by the NIHR (NIHR155654).

This review was conducted on behalf of the Digital Interventions for South Asians with Cardiometabolic Disease Study Consortium [34].

## Authors’ Contributions

The study concept was designed by ABanerjee, MR and LP. Interviews were carried out by MR and NK. Analysis was conducted by MR and NK with support from DS. MR wrote the original draft with review and edits by ABanerjee, NK and DS. Additional review was carried out by LP, ABlandford, PG, KK, SM, KP, HWWP, and MS for the Digital Interventions for South Asians with Cardiometabolic Disease Study consortium.

