## Supplemental 1 for "Healthcare professionals’ attitudes towards digital health interventions and perspectives on digital health inequalities: a qualitative study"

### Supplementary 1: Focus group guide for HCPs: barriers and facilitators to uptake of DHIs at an individual and intervention level

NB: Schedules were adapted for individual interviews and according to the professional composition of the group

| | |
|---|---|
| Study and participant introduction | • Welcome<br>• Permission to record<br>• Participant introductions<br>• Introduction of focus group aims and outline of the session<br>• Online session 'rules' |
| DHIs introduction | • Definition of DHIs<br>• Allow time for questions before start |
| *Part 1* | *In this section we seek to understand use of DHIs in South Asian populations, and for CVD and DM.* |
| Professional experience of DHIs for CV or DM | • What professional experience do you have with DHIs for cardiovascular disease or diabetes (or for other health problems)?<br>*Prompts:*<br>○ *E.g. working at front-line with patients on implementation and use, developing technology, or other?*<br>○ *If no, what is your professional experience related to DHIs or CMD?*<br><br>• In your experience, how and how often are DHIs used for CV/DM?<br>• Do these DHIs replace, enhance or provide additional care and support for patients? |
| Barriers and facilitators to use | In this section we will focus on barriers and facilitators to use. You can answer in reference to: DHIs you have experience of or other DHIs which have been used in cardiovascular disease, diabetes, or which affect these patient groups.<br><br>• In your opinion, what features of DHIs for CV or DM affect their accessibility and usability to patients and the public?<br>○ *What are the facilitators or* barriers?<br>○ What features affect how a DHI delivers its intended outcomes?<br><br>• As a health professional, what factors impact whether you recommend use of a particular DHI to a patient?<br>○ *For other HCPs, what is your experience/interactions with patients related to DHIs for CMD/CMD management?*<br>○ *Which apps have been more or less useful in the long-term?*<br><br>• In your experience, what other reasons are there for end users to want or not want to use DHIs? |
| Population differences in use of DHIs | <u>Wider population</u><br>• Are you aware of any patterns of differences in use of DHIs, for example by age, socioeconomic status, or ethnicity?<br>○ *What do you think are the reasons for these differences?*<br>○ *As use of DHIs becomes more widespread in health and care, which groups do you think will have greater or less access?* |

| | |
|---|---|
|  | <u>Population – South Asian</u><br>• Focussing on experience of DHIs of those of a South Asian background, can you share your experience or opinion on whether particular DHIs have been successful or unsuccessful in South Asian communities?<br> ○ *What about these made them successful/unsuccessful?*<br><br><u>Population - inequalities</u><br>• What types of alternatives can you offer those without digital access?<br>• Can you tell me about anything that could help people in these groups use DHIs (that we haven't already discussed)? |
| *Part 2* | *In this section we will explore the impact of the coronavirus pandemic on use of DHIs. Recognise that this is a sensitive topic for many individuals, and if people feel the need to withdraw at any point in the discussion they can do so.* |
| Coronavirus impact on health management | • How has the coronavirus pandemic changed the way you support patients?<br> ○ *to manage CVD/DM?*<br> ○ *changes in use to DHIs?* |
| Use of coronavirus DHIs/ Coronavirus and inequalities | • In your experience, how have patients and the public used technology to get information about coronavirus, or to take any action related to coronavirus (like testing or vaccination)?<br> ○ *If applicable to your area of work, were you able to offer official information about coronavirus or about action, such as testing or vaccination, in ways that are not digital?*<br> ○ *If specific information related to coronavirus and CVD/DM was available, were the formats appropriate to your patient population?*<br><br>• In your experience or opinion, have there been inequalities arising from the use of technology in healthcare during coronavirus?<br> ○ *Were there any specific patterns of use that you found interesting or concerning? (Any specific to South Asian populations?)* |
| Wrap-up | • Summarise key points in each topic<br>• Any new or closing comments from participants |
| Session close | • Thank for participation<br>• Signpost to consent form regarding consent, data protection, outputs of session and contacting research team<br>• Remind participants that they will receive their £50 voucher and a sources of further support sheet in the post. |
